# Role of asymptomatic COVID-19 cases in viral transmission: Findings from a hierarchical community contact network model^1^

**DOI:** 10.1101/2020.11.21.20236034

**Authors:** Tianyi Luo, Zhidong Cao, Yuejiao Wang, Daniel Dajun Zeng, Qingpeng Zhang

## Abstract

**Background:** As part of on-going efforts to contain the COVID-19 pandemic, understanding the role of asymptomatic patients in the transmission system is essential to infection control. However, optimal approach to risk assessment and management of asymptomatic cases remains unclear.

**Methods:** This study involved a SEINRHD epidemic propagation model, constructed based on epidemiological characteristics of COVID-19 in China, accounting for the heterogeneity of social network. We assessed epidemic control measures for asymptomatic cases on three dimensions. Impact of asymptomatic cases on epidemic propagation was examined based on the effective reproduction number, abnormally high transmission events, and type and structure of transmission.

**Results:** Management of asymptomatic cases can help flatten the infection curve. Tracking 75% of asymptomatic cases corresponds to an overall reduction in new cases by 34.3% (compared to tracking no asymptomatic cases). Regardless of population-wide measures, family transmission is higher than other types of transmission, accounting for an estimated 50% of all cases.

**Conclusions:** Asymptomatic case tracking has significant effect on epidemic progression. When timely and strong measures are taken for symptomatic cases, the overall epidemic is not sensitive to the implementation time of the measures for asymptomatic cases.

## Introduction

The severe acute respiratory syndrome coronavirus 2 is a type of coronavirus that has caused the pandemic known as the Coronavirus Disease of 2019 (COVID-19). It remains a major global health threat at the time of writing. By October 2020, there were >38 million confirmed cases worldwide. The outbreak of COVID-19 in China was under control in April. However, in June and October, two cluster infections of more than 100 people occurred in Beijing and Kashgar, China, respectively. In these two outbreaks, many patients were asymptomatic and were identified through close contact tracking and screening. Empirical studies have indicated that individuals may be most infectious during the presymptomatic phase (He et al., 2020). Undetected cases of asymptomatic infection may be an important source of infection, and symptom-based screening was insufficient to detect a high proportion of infectious cases (Day, 2020). Some experts (Qiu, 2020) speculate that 59% of early cases in Wuhan remained undiagnosed, including cases that remained asymptomatic or developed mild symptoms. Therefore, asymptomatic patients cannot be ignored in the chain of infection. Understanding the impact of presymptomatic phase or asymptomatic cases on COVID-19 transmission will be fundamental to the success of control strategies after the first outbreak (Moghadas et al., 2020). The effectiveness of symptom-based interventions depends on the proportion of asymptomatic infections, the infectiousness of asymptomatic cases, and the duration and infectiousness of the presymptomatic phase. These have caused significant social concern, in particular, regarding the risk of another outbreak caused by asymptomatic cases. These worries are well founded, as the role of asymptomatic cases in COVID-19 spread remains unclear and the most optimum approach to asymptomatic case management has not been elucidated. As a result, these two questions have attracted significant research interest.

Since April 1, the Chinese authorities have been publishing daily figures on asymptomatic coronavirus cases, suspecting that asymptomatic cases were driving epidemic spread. At the time of writing, there have been many case studies and epidemiological studies based on asymptomatic cases (Bai et al., 2020; Hu et al., 2020). Empirical studies (Mizumoto et al., 2020; Nishiura et al., 2020) indicate that asymptomatic infections account for 17.9 to 30.8% of all infections. Some studies (Anastassopoulou et al., 2020; Eikenberry et al., 2020; Panovska-Griffiths et al., 2020) considered asymptomatic patients in the process of epidemic spread modeling, but few studies focus on the evaluation of interventions for asymptomatic patients and the impact of asymptomatic patients on the second outbreaks (Ali et al., 2020; Wang et al., 2020). The role of asymptomatic cases in the transmission chain remains unclear.

Understanding the role of asymptomatic cases in infection spread is critical to the prevention and effective management of future outbreaks. In transition to long-term management of COVID-19, understanding the role of asymptomatic cases can inform public health policies. To quantify the effect of interventions on asymptomatic patients in community prevention and control, we researched the impact of asymptomatic case tracking on the spread of the epidemic based on a community-level social network. Based on 40 empirical studies on COVID-19, we determined the epidemiological characteristics of COVID-19 in China, such as the proportion of asymptomatic patients and its infectiousness. We considered the structure of the local community and social network, on which we proposed a SEINRHD propagation model that uses a hierarchical community network. We used this model to conduct computational experiments that evaluated the impact of asymptomatic case management on infection curves in three dimensions. We explored the impact of asymptomatic case tracking ratio, diagnosis delay time, and strategy implementation timing on epidemic progression. Finally, we examined the propagation characteristics of a benchmark and two alternative scenarios, including asymptomatic case tracking impact on the effective reproduction number, type of transmission link, and abnormally high transmission events. Our results highlight the need for timely implementation of strategies for asymptomatic patients (such as contact tracing) in community prevention and control and the need for family isolation. The examined interventions can help flatten the new infection curve.

## Methods

### Generation of the hierarchical community contact network

A hierarchical network was constructed, representing social contacts within the Chinese community. All network nodes belonged to a big community, which was divided into seven small communities. Nodes within each community represented connections between individuals who lived in geographical proximity and shared characteristics such as age or interests. A small community comprised multiple households and constituted a fully connected network at the bottom of hierarchical community network. Social contacts within this network had a hierarchical community structure. One of the contacts was associated with the highest risk of infection acquired from a family member, with a moderate risk of infection acquired from a member of another household within the same community, and a low risk of infection acquired from members of small diverse communities. In addition, the network included community workers that had frequent contact with all small communities. All ties within a network were defined as undirected. The number of individuals in the network was denoted by *n*. In our simulation, n=10,000. A schematic of the network structure is presented in figure 1. The network was created in the following process. First, the number of small groups was determined. The number of big community nodes was set to 10,000, of which 200 were community workers. The remaining 9,800 nodes were divided into 7 small communities. The number of people within these seven communities was subject to Poisson distribution. Each small community consisted of families; the number of families followed a Poisson distribution with an average value of four.

**Figure 1.**
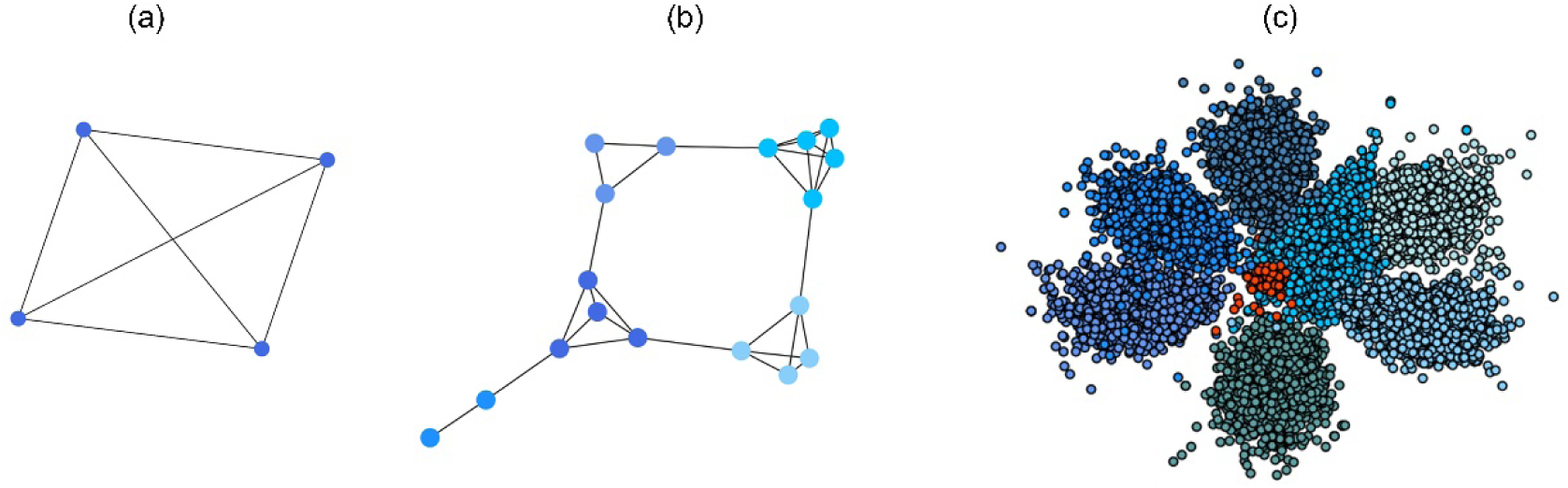
(a) Schematic of a household-based social network, whereby all family members are connected. (b) Schematic of a small community social contact network, where different colored nodes represent different households, showing many connections within each household, and fewer connections between households. (c) Schematic of a hierarchical network, where a big community is divided into smaller communities, represented by different color nodes. Some edges exist in each small community, with fewer edges connecting small communities. The red nodes represent community workers, which is a special group within the big community.

Second, between-node connections were determined. Within a household, all nodes were connected to each other. A household constituted a fully connected network. Nodes within the same small community but part of different households were interconnected with a probability of 0.001. Nodes belonging to different small communities were interlinked with a probability of 0.000001. Social connections of 200 community workers were represented by a small-world network. The average degree of this network is 25.

### Transmission model

We modeled the spread of COVID-19 using a compartmental epidemic model SEINRHD. This model is an extension of the SEIR model and is adapted to the China context. Considering the spread and treatment strategies of COVID-19 in the Chinese environment, we have added asymptomatic status, hospitalized and reported, and death status and classified symptomatic status according to the severity of the disease.

The model of transmission dynamics for influenza pandemics classifies individuals as susceptible (S); exposed (E); clinically ill and infectious (I), infectious individuals are divided into either asymptomatic or in different symptomatic groups: mild, severe, or critical symptoms; hospitalized and reported (H); recovered (R); and death (D) (figure 2). The default parameters of this model are determined by the early epidemiological characteristics of COVID-19 in China. More detailed model parameter settings will be explained in the next section.

**Figure 2.**
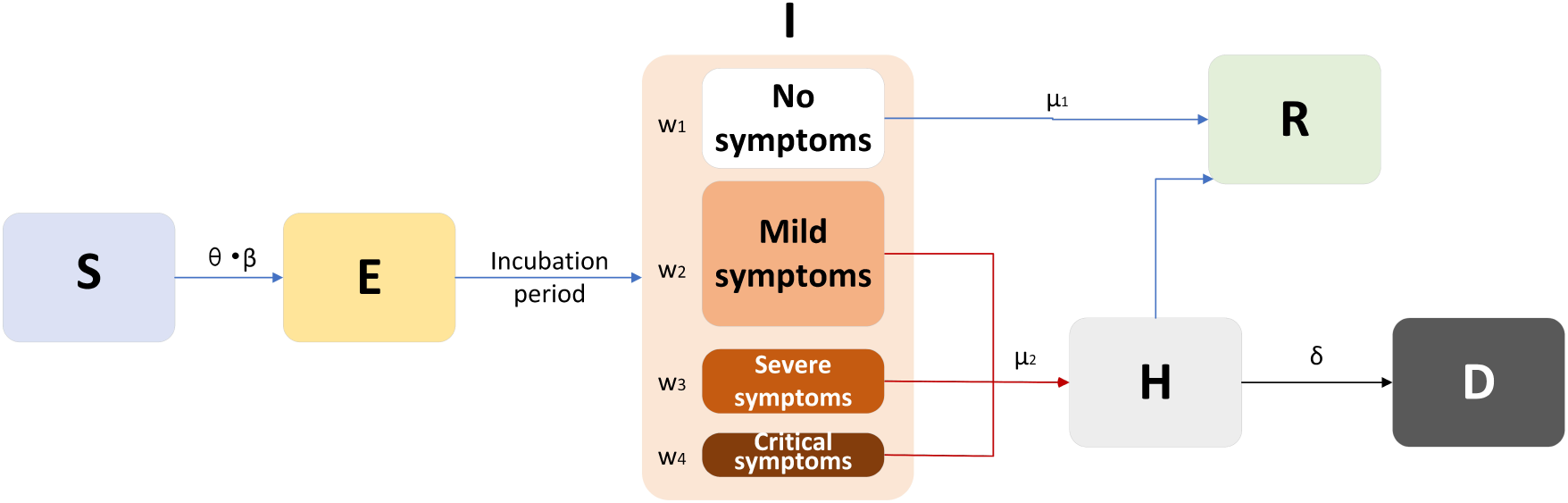
The SEINRHD model of epidemic progress.

Transmission occurs through social interactions. A schematic diagram of the SEINRHD propagation model is shown in figure 2. The model comprises the following steps:

1. Within the network, a seed node is randomly selected, designated as E, and the remaining nodes become S.
2. Node E becomes I, following an incubation period of *T*_*e*_. The node is considered infectious during state I state and on the last day of state E.
3. Throughout the process, the infectious node infects its susceptible neighbors with a probability expressed as *θ*. *β*.
4. There are four types of patients with I status. Among them, asymptomatic patients become recovered after an average period of *μ*_1_ days. The rest of the infected nodes are hospitalized and diagnosed within an average delay of *μ*_2_ days.
5. The mortality rate among hospitalized nodes is *δ*; the remaining cases recover after a period of treatment.
6. The process ends once there is no node exposed or infectious.

We defined *θ* as the probability of contact between individuals and *β* as the probability of infection after contact. *θ*. *β* represented disease transmission rate between nodes.

### Experimental parameters

To make the initial value of the model parameters conform to the epidemiological characteristics of COVID-19, we performed a literature review of 36 related studies, summarizing dynamic and symptomatic characteristics of COVID-19. First, we estimated the average number of mild, severe, and critical symptomatic cases. (Guan et al., 2020; D. Wang et al., 2020; Wu & McGoogan, 2020), (http://wjw.sz.gov.cn/). At the early stages of the epidemic, when knowledge about the asymptomatic cases was limited, most studies only reported cases that progressed to mild, severe, and critical symptoms. Second, we estimated the proportion of asymptomatic cases as 27.3% (Kimball et al., 2020; Mizumoto et al., 2020; Qiu, 2020, Chen et al., 2020). Using these two sets of values to estimate the proportion of four types patients in the model. Table 1 presents model parameters on benchmark scenario (model parameters consistent with COVID-19 characteristics reported early in the pandemic and prior to any interventions.). The incubation period is a characteristic of infectious disease. Our literature review of studies on COVID-19 incubation period included 17 articles (see Supplementary Table 1), yielding an estimated value of 5.11 days, with the upper bound of 72% <10.2. The incubation period in our model followed a normal distribution with a mean of 5.11 and a variance of 2.5. Symptomatic infection period *μ*_2_, asymptomatic infection period (days) *μ*_1_, and mortality rate *δ* were estimated based on findings from early studies of COVID-19 (Guan et al., 2020; Liu et al., 2020; Sun et al., 2020; D. Wang et al., 2020; K. Wang et al., 2020; Wu & McGoogan, 2020). *θ*. *β* represented disease ability to spread within a population. We used the basic reproduction number *R*_0_ = 3.11, which is based on early analysis of Wuhan COVID-19 data (Read et al., 2020), to estimate the propagation rate in our model. The reproduction number *R*_0_ is defined as the average number of new infections generated by one infected individual during the entire infectious period in a fully susceptible population. The method of calculating *R*_0_ in our model see Supplementary).

**Table 1.** Model parameters.

| Variable | Meaning | Value or distribution |
| --- | --- | --- |
| $w_1$ | Proportion of asymptomatic patients | 27.3% |
| $w_2$ | Proportion of mildly symptomatic patients | 55.9% |
| $w_3$ | Proportion of severely symptomatic patients | 10.0% |
| $w_4$ | Proportion of critically symptomatic patients | 6.8% |
| $T_e$ | Incubation period (days) | N (5.11, 2.5) |
| $\mu_2$ | Symptomatic infection period (days) | 7 |
| $\mu_1$ | Asymptomatic infection period (days) | 7.5 |
| $\delta$ | Mortality rate | 3.63% |
| $\theta \cdot \beta$ | Propagation rate | 0.036 |

In the strategy evaluation experiment, measures taken for asymptomatic patients included increasing the tracking range and accelerating the detection time. In all experiments, the same measures were taken for symptomatic patients: when the diagnosed (reported) case number was 10, the delay in diagnosis of symptomatic patients reduced from 7 to 3 days. Experiment parameters are presented in Table 2, including ρ, which represented the tracking proportion of asymptomatic patients. *T*_*i*_ represented time to strategy implementation, which was triggered when the cumulative number of confirmed cases exceeded a defined threshold.

**Table 2.** Model parameters per experiment.

| Experiment 1: |  | Experiment 2: |  | Experiment 3: |  |
| --- | --- | --- | --- | --- | --- |
| $\mu_1 = 3$ , $\mu_2 = 3$ , $T_i =$<br>*10 reported* | | $\rho = 75\%$, $\mu_2 = 3$, $T_i =$<br>*10 reported* | | $\rho = 75\%$, $\mu_1 = 3$, $\mu_2 = 3$ | |
| 1 | $\rho = 0$ | 1 | $\mu_1 = 3$ | 1 | $T_i = 10$ *reported* |
| 2 | $\rho = 25\%$ | 2 | $\mu_1 = 4$ | 2 | $T_i = 40$ *reported* |
| 3 | $\rho = 50\%$ | 3 | $\mu_1 = 5$ | 3 | $T_i = 70$ *reported* |
| 4 | $\rho = 75\%$ | 4 | $\mu_1 = 6$ | 4 | $T_i = 100$ *reported* |
| 5 | $\rho = 100\%$ | 5 | $\mu_1 = 7$ | | |

Experiment 1 examined the effect on disease spread of tracking rate of the asymptomatic cases. Experiment 2 investigated the effect of delayed diagnosis time on asymptomatic patients on the epidemic. Experiment 3 studied the impact of the time to implement the strategy of tracking asymptomatic patients on the epidemic. The results of each group of experiments are the statistical results after 1000 simulations under this parameter setting. The total duration of all experiments is 210 days. In the first four sections of the results, the starting point of the abscissa time is when the number of confirmed reports is 10 (day 31).

### Characteristics of the transmission

To better study the changes of various propagation characteristics after taking measures, in addition to the above three strategy evaluation experiments, we focused on the analysis of the transmission characteristics in the following two situations: only taking measures for symptomatic patients (*ρ* = 0, *μ*_1_ = 3, *μ*_2_ = 3, *T*_*i*_ = 10 *reported*) and taking measures for symptomatic and asymptomatic patients at the same time (*ρ* = 75%, *μ*_1_ = 3, *μ*_2_ = 3, *T*_*i*_ = 10 *reported*) and conducted a comparative analysis with the benchmark scenario.

The following are some terms and their definitions involved in describing transmission characteristics.

Effective reproduction number *R*_*e*_ : The basic reproduction number *R*_0_ is defined as the average number of secondary infections caused by a typical primary infection in a fully susceptible population. *R*_0_ is one of the most important epidemiological parameters when monitoring an epidemic because it is fundamental to assess the potential spread of the virus. Its value changes during an epidemic, and it is termed as the effective reproduction number *R*_*e*_. *R*_*e*_ ; it can be used to observe the control of infectious diseases, especially whether the government can reduce *R*_*e*_ to below 1, or even to a very low level through prevention and control measures.

### Degree

In the transmission tree graph, the degree of a node indicates the number of connections of the node. Among these connections, there is a node that infects disease to this node, called the parent node. The rest of the nodes are infected by the node, called child nodes.

Complementary cumulative distribution function (CCDF): CCDF can completely describe the probability distribution of a variable *a*. CCDF represents the sum of the occurrence probability of all values greater than *aa* for a continuous function:

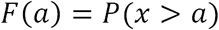

## Results

### Impact of asymptomatic case tracking on disease spread

In experiment 1, with fixed parameters (*μ*_1_ = 3, *μ*_2_ = 3, *T*_*i*_ = 10 *reported*), the tracking rate of asymptomatic cases was 0, 25%, 50%, 75%, and 100%, respectively. Figure 3 presents epidemic progression under different scenarios.

**Figure 3.**
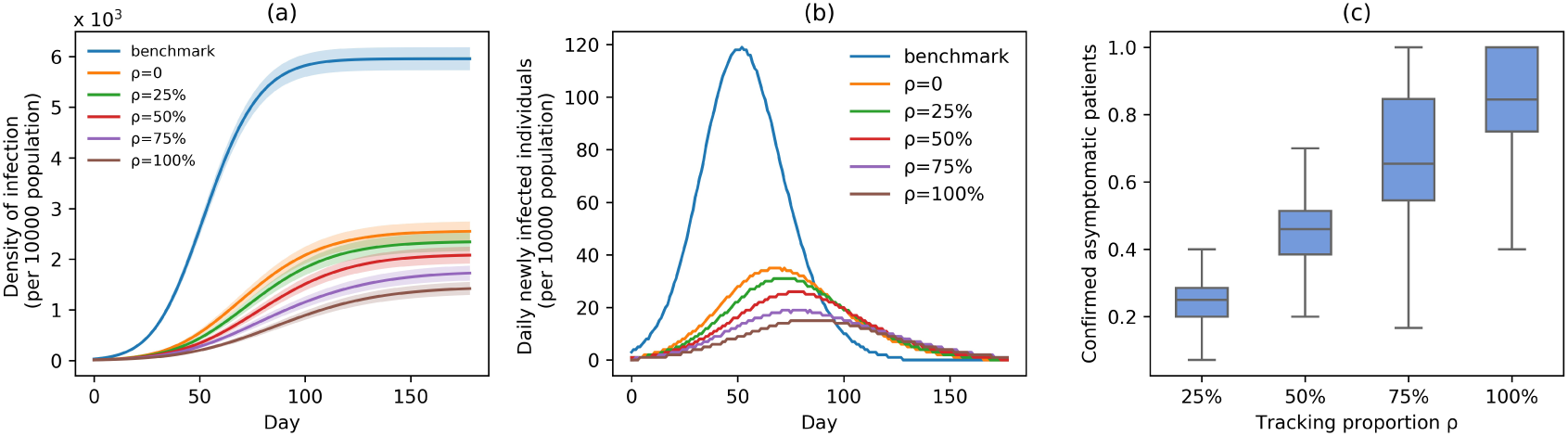
Infection status under experiment 1 (asymptomatic patients at different tracking rates). (a) Changes in density of infection. The lines represent the mean density of infection per 10000 people, while the shaded areas represent the 95% reference range. (b) The number of daily newly infected individuals per 10000 people. (c) The proportion of confirmed asymptomatic patients in total asymptomatic patients under different schemes. Boxplots represent percentiles 2.5%, 25%, 50%, 75%, and 97.5% of the distribution.

Relative to the benchmark, control measures significantly impacted disease spread, resulting in infection density reduction inversely proportional to the number of asymptomatic cases identified. At day 100, relative to benchmark, infection density was reduced by 63.2% (*ρ* = 0), 67.4% (*ρ* = 25%), 72.9% (*ρ* = 50%), 79.2% (*ρ* = 75%), and 83.8% (*ρ* = 100%) (figure 3(a)). On the 150th day, the outbreak was nearly extinguished; at that stage, compared with benchmark, infection density was reduced by 3,438 (ρ=0), 3,652 (ρ=25%), 3,924 (ρ=50%), 4,304 (ρ=75%), and 4,605 (ρ=100%) per 10,000 people (figure 3(a)). Overall, tracking 75% of asymptomatic cases corresponded to an outbreak reduction of 34.3%, compared to no tracking. Compared with the benchmark scenario, slowing disease spread associated with a 24.7-day delay in infection peak was associated with a reduction to the peak number of infections of 78.5% in scheme 3 (*ρ* = 50%). Compared with scheme 1(*ρ* = 0), slowing disease spread associated with a 10-day delay in infection peak was associated with a reduction to the peak number of infections of 25.7% in scheme 3 (*ρ* = 50%) (figure 3(b)). The proportion of asymptomatic patients (median) detected is slightly lower than the proportion set in our strategic plan (figure 3(c)).

### Impact of delayed asymptomatic patient diagnosis on epidemic progression

In experiment 2, at fixed parameters (*ρ* = 75%, *μ*_2_ = 3, *T*_*i*_ = 10 *reported*), the delay in asymptomatic patient diagnosis was 3, 4, 5, 6, and 7 days, respectively (figure 4).

**Figure 4.**
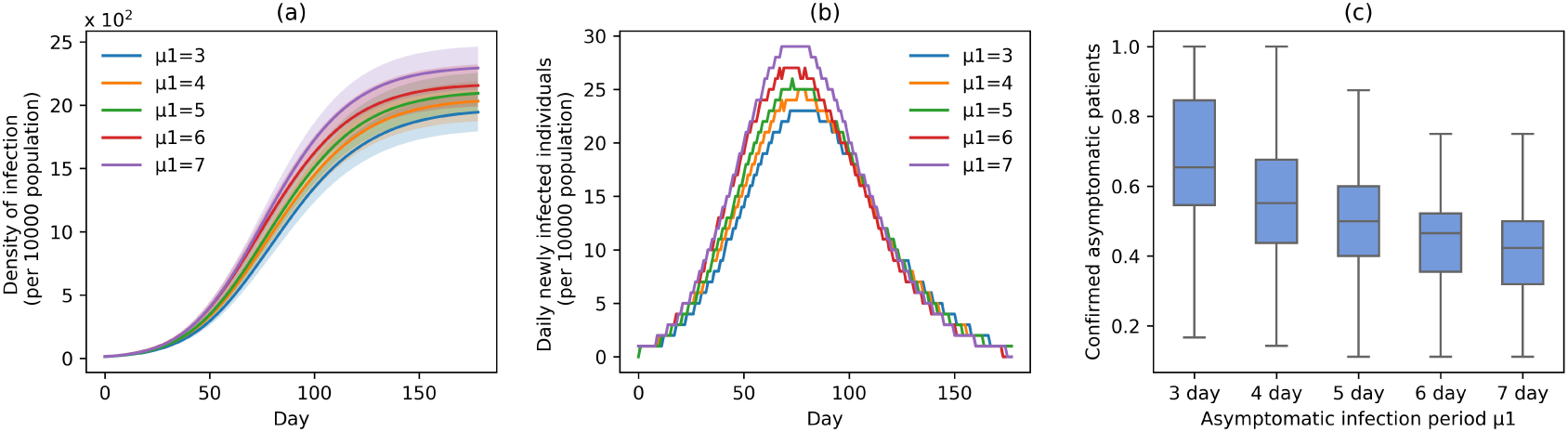
Infection status under experiment 2 (different delayed diagnosis time on asymptomatic patients). (a) Changes in density of infection. The lines represent the mean density of infection per 10000 people while the shaded areas represent the 95% reference range. (b) The number of daily newly infected individuals per 10000 people. (c) The proportion of confirmed asymptomatic patients in the total asymptomatic patients under different schemes. Boxplots represent percentiles 2.5%, 25%, 50%, 75%, and 97.5% of the distribution.

At increased speed of diagnosis, the epidemic appeared to come under control more quickly (figure 4(a)). At day 100, compared with scheme 5 (*μ*_1_ = 7), infection density was reduced by 21.2% (*μ*_1_ = 3), 15.6% (*μ*_1_ = 4), 12.3% (*μ*_1_ = 5), and 6% (*μ*_1_ = 6) (figure 4(a)). Compared with scheme 5 (*μ*_1_ = 7), the average infection curve peak delay was 5 days, which corresponded to a decrease in peak height of 20.7% in scheme 1 (*μ*_1_ = 3) (figure 4(b)). The peak count of daily new cases was 23 (*μ*_1_ = 3), 25 (*μ*_1_ = 4), 26 (*μ*_1_ = 5), 27 (*μ*_1_ = 6), and 29 (*μ*_1_ = 7) (figure 4(b)). The proportion of asymptomatic cases (median) detected at the end of the simulation was 65.4% (*μ*_1_ = 3), 55.2% (*μ*_1_ = 4), 50.0% (*μ*_1_ = 5), 46.6% (*μ*_1_ = 6), and 42.2% (*μ*_1_ = 7) (figure 4(c)).

### Impact of measure implementation timing on epidemic progression

Timing of measure implementation was expressed as *TT*_*ii*_. In experiment 3, with fixed parameters (*ρ* = 75%, *μ*_1_ = 3, *μ*_2_ = 3), when the number of confirmed cases reached 10, 40, 70, and 100, simulation interventions were implemented (figure 5). Under constant intervention intensity, timing of intervention implementation did not affect epidemic progression (figure 5(a) (b)), with the average number of new cases peaked around day 79 in all scenarios (figure 5 (b)).

**Figure 5.**
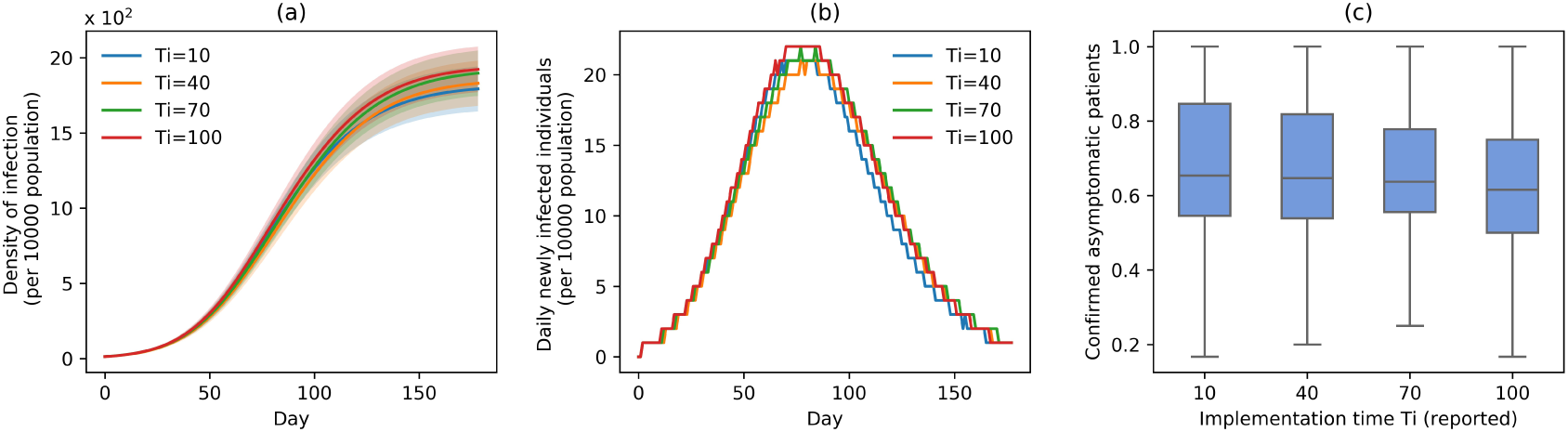
Infection status under experiment 3 (take measures for asymptomatic patients at different times). (a) Changes in density of infection. The lines represent the mean density of infection per 10000 people while the shaded areas represent the 95% reference range. (b) The number of daily newly infected individuals per 10000 people. (c) The proportion of confirmed asymptomatic patients in the total asymptomatic patients under different schemes. Boxplots represent percentiles 2.5%, 25%, 50%, 75%, and 97.5% of the distribution.

The proportion of asymptomatic cases (median) confirmed at the end of simulation was 66.7% (*TT*_*ii*_ = 10), 66.7% (*TT*_*ii*_ = 40), 66.7% (*TT*_*ii*_ = 70), and 61.5% (*TT*_*ii*_ = 100) (figure 5(c)). The mean proportion of asymptomatic cases (median) confirmed at the end of the simulation was 69.2% (*TT*_*ii*_ = 10), 68.5% (*TT*_*ii*_ = 40), 67.7% (*TT*_*ii*_ = 70), and 61.5% (*TT*_*ii*_ = 100) (figure 5(c)).

### Effective reproduction number

Figure 6 shows changes to *R*_*e*_ throughout the disease propagation period. Under the benchmark scenario, *R*_*e*_ was <1 after day 47. On days 0 to 25, *R*_*e*_ exceeded 2. *R*_*e*_ peaked at 3.63. Following the introduction of interventions, *R*_*e*_ was smaller than that estimated under the benchmark scenario. After 42 days, *R*_*e*_ in all three scenarios was <1 and gradually decreased further. Compared with the implementation of measures only for symptomatic patients, *R*_*e*_ is significantly reduced before the 42^nd^ day after adding the tracking strategy for asymptomatic patients.

**Figure 6.**
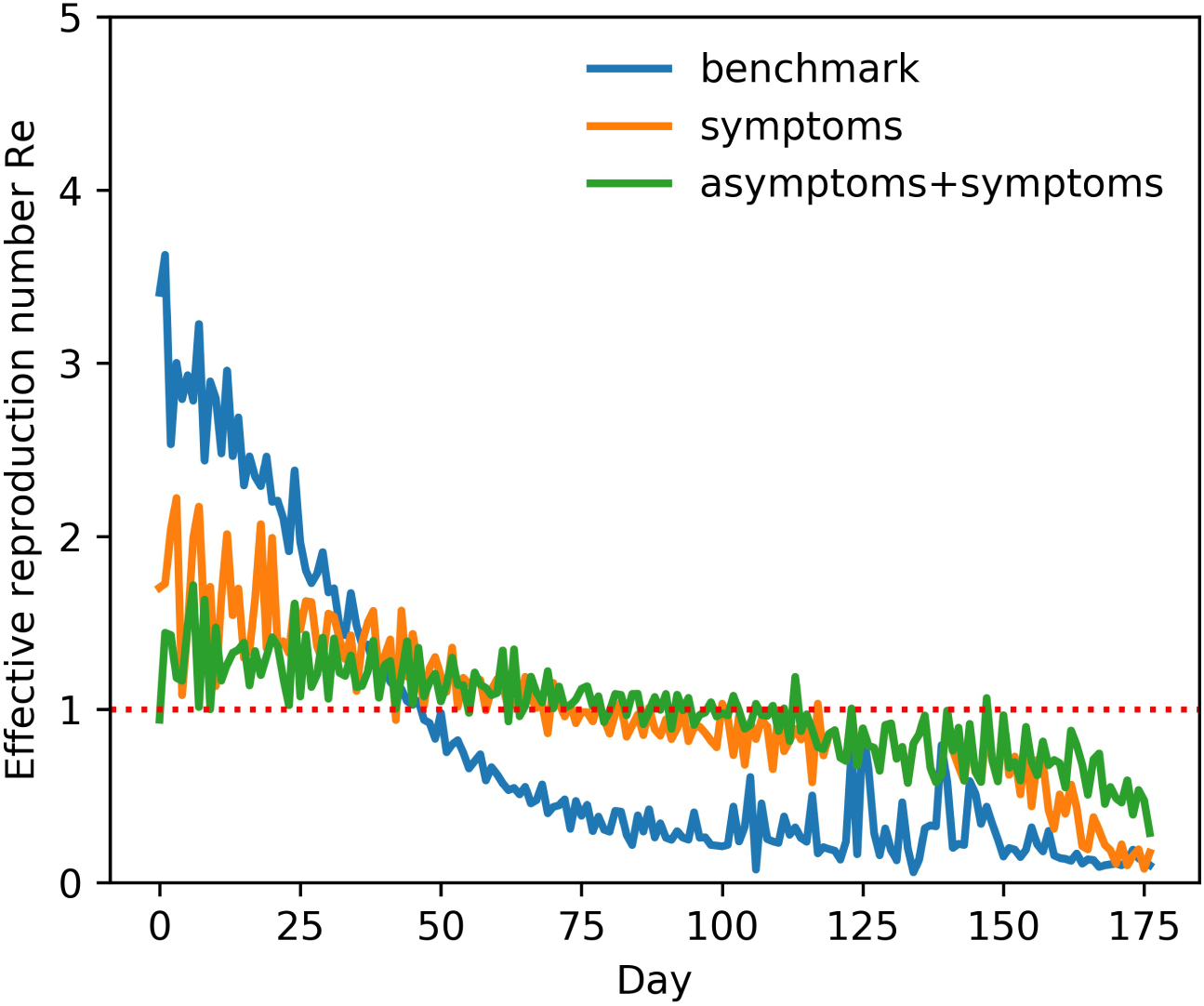
Effective reproduction number *Re* (mean) over time. Only take the control strategies for symptomatic patients (orange line), take the control strategies for both symptomatic patients and symptomatic patients (green line), benchmark scenario (blue line).

### Characteristics of infected individuals

Figure 7(a) shows time-dependent changes to the number of new cases within the network. As shown in figure 7(a), in the benchmark scenario, the virus preferentially infects nodes with larger degrees in the network, and then gradually infects the nodes with fewer degrees. Note that the abscissa of this figure corresponds to the time of first case confirmation. Following the implementation of interventions, viral transmission within the network slowed down significantly. It suppresses the infection of the virus to the nodes with the larger degree. As the virus spreads, the degree distribution of newly infected nodes is relatively uniform. In addition, in the early stages of disease spread, the average degree of newly infected nodes showed strong oscillations. Tracking asymptomatic cases reduced the average degree of newly added nodes after day 50, compared to tracking only symptomatic patients.

**Figure 7.**
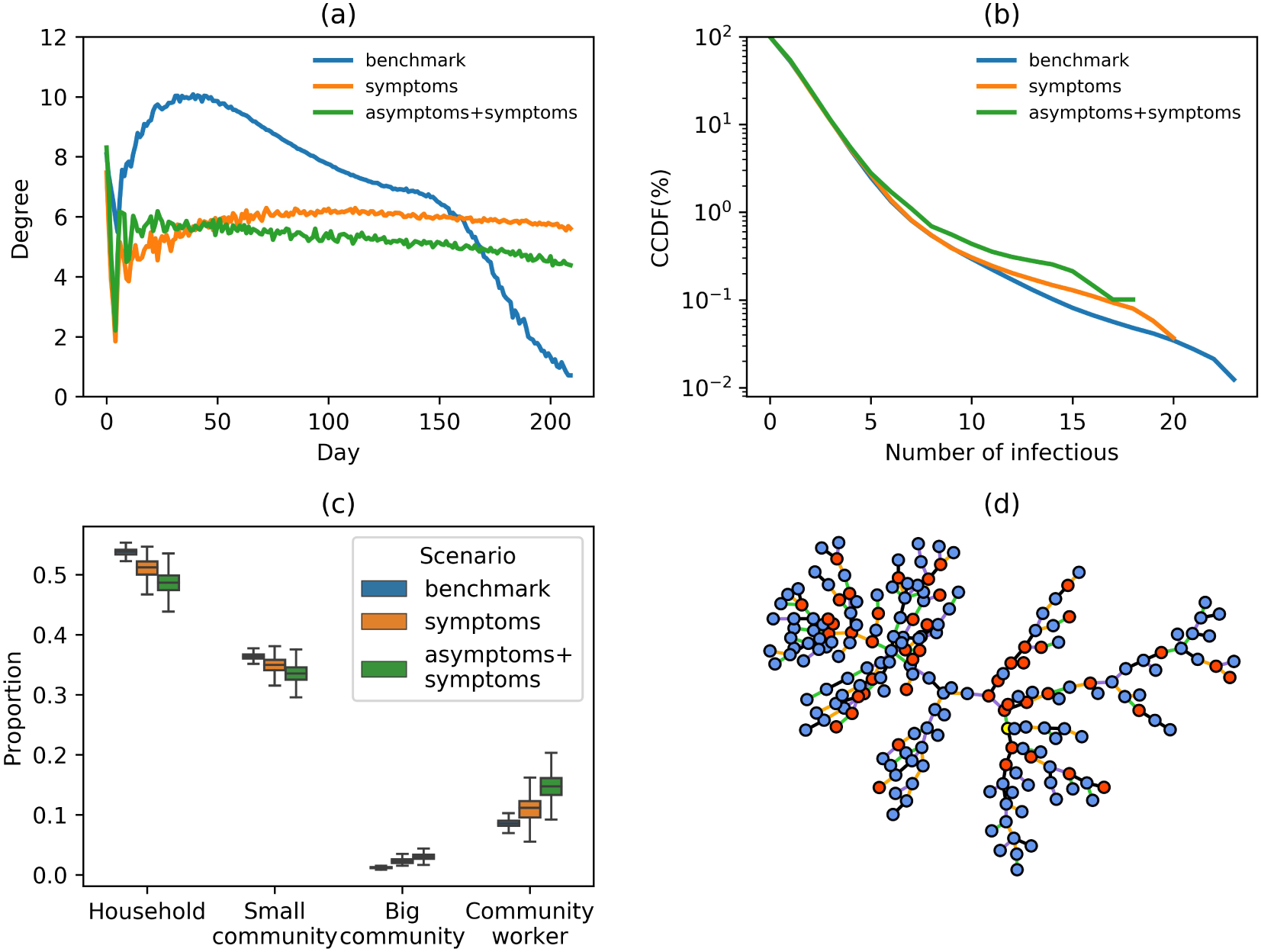
(a) Changes to the proportion of newly infected nodes within a network over time. (b) Complementary cumulative distribution function (CCDF) of the number of secondary infections per infected individual. Only take the control strategies for symptomatic patients (orange line), take the control strategies for both symptomatic patients and symptomatic patients (green line), benchmark scenario (blue line). (c) The proportion of transmission types (infected by household, small community, big community or community workers). (d) Example of a transmission tree in a simulation experiment. (color of the nodes, yellow, first case; red, asymptomatic infection; blue, symptomatic infection; color of the lines for the type of transmission, black, among household members; green, in the small community; orange, in the big community; purple, infected by community workers).

Figure 7(b) shows the CCDF of the number of secondary infections per individual. Before and after interventions, approximately 75% of cases infected one or two people. In the benchmark scenario, the maximum number of secondary infections caused by a single case reached 25; approximately 0.04% of primary cases corresponded to over 20 secondary cases. After introducing tracking of symptomatic and asymptomatic cases, the maximum number of secondary infections per single primary case was reduced to 18. When interventions were applied only to symptomatic patients, the maximum number of secondary infections per single primary case was 21. When interventions were also applied to symptomatic patients, abnormally high transmission events were reduced. Notably, when interventions included asymptomatic cases, abnormally high transmission events reduced further.

### Characteristics of the transmission tree

Figure 7(c) shows the proportion of transmission types. Figure 7(d) presents an example of a transmission tree in a simulation experiment. Under all intervention scenarios, approximately half of all new cases were infected by family members (figure 7(c)), suggesting a necessity to reduce social contacts in small communities and strengthen the protection of community workers. Our model captured the back-and-forth transmission patterns between households, small communities, and community workers, as shown in figure 7(d). Furthermore, asymptomatic cases seem to play a role in the transmission chain.

## Discussion

In the absence of a vaccine against COVID-19, governments and organizations face economic and social pressures to gradually and safely lift social distancing measures. To prevent epidemic rebound during long-term epidemic management, it is vital to understand the role of asymptomatic cases in disease transmission. The present study examined the epidemiological characteristics of COVID-19 and hierarchical characteristics of the Chinese community contact network to assess the impact of asymptomatic cases on three dimensions of disease transmission, aiming to provide evidence for future decision making.

The results show that tracking a proportion of asymptomatic cases, detection strength of asymptomatic cases, and timing of asymptomatic case tracking can reduce the cumulative number of disease cases.

The examined interventions can help flatten the new infection curve. Increasing the proportion of asymptomatic cases being tracked can have the most significant impact on disease spread. However, after fixing the intensity of the detection of symptoms and the strength of asymptomatic measures, the overall epidemic situation is not sensitive to the implementation time of the measures. With the same intensity of interventions, the implementation of the measures after two months has little effect on the density of infection.

From the perspective of transmission characteristics, after taking measures for symptomatic patients, the number of viruses regenerating rapidly decreased, and the ability to spread significantly weakened. Concurrently, primary cases associated with the highest number of secondary cases can be effectively contained. This combination of strategies can help reduce the rate of viral transmission and ultimately extinguish the epidemic. These measures can also reduce the risk of occurrence of super-spreaders.

This study also examined characteristics of a transmission link, showing that, in the absence of interventions, within-family transmission accounts for nearly half of new cases, while transmission rate within large communities remains within 3%.

The present study findings can inform public health policy regarding asymptomatic cases of COVID-19 worldwide. First, the most important aspect of a strategy involving asymptomatic case control is the tracking ratio. Therefore, in actual prevention and control, measures to track and isolate the asymptomatic cases are useful. Given human and economic resource restriction, reducing the number of tracked cases will be unavoidable, leading to small fluctuations in the number of new confirmed cases; however, the epidemic can still be effectively controlled. Second, in the early stages of a pandemic, quarantine of symptomatic patients should be prioritized to achieve early detection, rapid isolation, and timely treatment. Given insufficient medical and socioeconomic resources, interventions aimed at asymptomatic patients can be introduced strictly in the second phase of epidemic control, when the initial outbreak has been contained. Third, as household transmission accounts for half of new cases, it should be valued by the general public and relevant departments. Disease control and prevention within families should be emphasized during an epidemic. Community workers play a critical role in disease spread within large communities, suggesting these teams should be equipped in personal protection gear to curtail their role in the transmission chain.

To date, most studies examining the role of asymptomatic cases on disease transmission have involved small sample sizes. Combined with differences in definitions of asymptomatic cases between countries, these studies have reported inconsistent findings. In particular, the number of asymptomatic cases and the amount of time a person with COVID-19 remains a carrier is still unclear. Our model can help estimate the risk of another COVID-19 wave and evaluate realistic control and prevention strategies. At the start of a second wave in Beijing, China, which occurred around June 11, rapid case and contact tracking and isolation, combined with large-scale testing, including among suspected but asymptomatic cases, helped prevent a sizeable outbreak.

This study has several limitations. First, asymptomatic cases considered in our model were cases that remained asymptomatic throughout the infection period. An alternative definition of “asymptomatic” refers to remaining symptom-free during the incubation period alone. To the best of our knowledge, no studies have compared prevention strategies applicable to these different categories of asymptomatic cases.

In summary, our model provides individuals, governments, and organizations with strategic insights for the management of asymptomatic cases during a pandemic. This study provides suggestions on intervention implementation, including priority, intensity, and target population. It has specific significance for alleviating the strict blockade measures and the social, medical, and economic burdens.

## Data Availability

The data that support the findings of this study are available from the supplemental material and it can be open accessed.

## Acknowledgments

This study was funded by National Natural Science Foundation of China (Nos. 72042018,91546112, 71621002) and Beijing Municipal Natural Science Foundation (No. L192012).

## ^1^Abbreviations

COVID-19: Coronavirus Disease of 2019.

