## Supplementary for "Role of asymptomatic COVID-19 cases in viral transmission: Findings from a hierarchical community contact network model^1^"

**Table 1. Estimated incubation period from selected studies.**

| **Author** | **Incubation period** | **Uncertainty** | **Source** |
|---|---|---|---|
| Mike(Mike, 2020) | 5.40 | 4.20-6.70 (95% CI) | NA |
| Liu et al. (Liu et al., 2020) | 4.80 | 2.20-7.40 | bioRxiv |
| Akhmetzhanov et al. (Akhmetzhanov et al., 2020) | 5.10 | 2.00-14.00 (95% CI) | medRxiv |
| Sun et al.(Sun, Chen, & Viboud, 2020) | 4.50 | 3.00–5.50 (IQRs) | The Lancet Digital Health |
| Read et al.(Read, Bridgen, Cummings, Ho, & Jewell, 2020) | 3.00 | 0.00-6.00 | medRxiv |
| Li et al.(Hui et al., 2020) | 5.20 | 4.10-7.00 (95% CI) | NEJM |
| Guan et al.(W.-j. Guan et al., 2020) | 3.00 | 0.00-24.00 | medRxiv |
| Sanche et al.(Sanche et al., 2020) | 4.20 | 3.50-5.10 (95% CI) | medRxiv |
| Zhang et al.(Zhang et al., 2020) | 5.20 | 1.80-12.40 | medRxiv |
| Tindale et al.(Tindale et | 9.00 | 7.92-10.20 (95% CI) | medRxiv |

| | | | |
|---|---|---|---|
| al., 2020) | | | |
| Ping(Ping, 2020) | 8.06 | 6.89-9.36 (95% CI) | medRxiv |
| Jing et al.(Jing et al., 2020) | 8.13 | 7.37-8.91 (95% CI) | medRxiv |
| Li et al.(Li et al., 2020) | 3.69 | 3.28-4.03 (95% CI) | Science |
| Li et al.(Li et al., 2020) | 3.6 | 3.41-3.91 (95% CI) | Science |
| Li et al.(Li et al., 2020) | 3.44 | 3.26-4.06 (95% CI) | Science |
| Bi et al.(Bi et al., 2020) | 4.80 | 4.20-5.40 (95% CI) | medRxiv |
| Guan et al.(W. Guan et al., 2020) | 4.00 | 2.00-7.00 (IQRs) | NEJM |
| Lauer et al.(Lauer et al., 2020) | 5.20 | 4.40-6.00 (95% CI) | medRxiv |
| Backer et al.(Backer, Klinkenberg, & Wallinga, 2020) | 6.40 | 5.60-7.70 (95% CI) | EuroSurveill |

### Calculation of $R_0$ in the model

Mathematical properties: For a non-negative stochastic variable $a$ that is distributed according to a probability density function $g(a)$, the moment generating function is defined as $M_{g(a)}(z) = \int_{a=0}^{\infty} e^{za}\ g(a)\mathrm{d}a$. When the integral is finite, there is a unique relation between the probability density function and its moment generating function. The moment generating functions have a number of mathematical properties.

In the paper (Wallinga & Lipsitch, 2007) have exploited the following properties: When the random variable $a$, distributed according to $g(a)$, can be thought of as the sum of two independent random variables $a1$ and $a2$, such that $a = a1 + a2$, with $a1$ and $a2$ distributed according to $f(a1)$ and $h(a2)$, the moment generating function for the random variable a can be composed of the moment generating function for $a1$ and $a2$: $M_{g(a)}(z) = M_{f(a)}(z) \times M_{h(a)}(z)$.

The moment generating function for the generation interval can be composed from moment generating functions for the durations of successive stages in the infection cycle. We introduce two distributions for the successive stages: $f(a)$ gives the duration from infection to becoming infectious, and $h(a)$ gives the duration from becoming infectious to infection. The moment generating function for the composite generation interval is then given by:

$$M_{g(a)} = M_{f(a)} \times M_{h(a)}$$

In our model, the incubation period obeys the normal distribution, and the infectious period obeys the exponential distribution.

$$R = \frac{1}{M(z)} = \frac{1}{M_{f(a)}(z)} \cdot \frac{1}{M_{h(a)}(z)}$$

$\frac{1}{M_{f(a)}(z)} = e^{\mu r - \frac{1}{2}\sigma^2 r^2}$ [f(a) incubation period obeys the normal distribution]

$\frac{1}{M_{h(a)}(z)} = (1 + \frac{r}{b})$ [h(a) infectious period obeys the exponential distribution]

Therefore, $R = e^{\mu r - \frac{1}{2}\sigma^2 r^2} \cdot (1 + \frac{r}{b})$.
